# Cardiopulmonary resuscitation (CPR) competency retention among registered nurses in critical care versus general care unit

**DOI:** 10.1101/2023.10.16.23297121

**Authors:** Yahia AL-Helih, Majeda AL-Ruzzieh, Sami AL-Yatim, Mohammad Alawneh, Saleh Abu-AL Haija, Faten Odeh

## Abstract

**Background:** Cardiac arrest is a sudden and life-threatening event resulting in an end of cardiac activity, necessitating immediate intervention to prevent fatalities. In-hospital cardiac arrest (IHCA) presents a critical medical emergency, demanding swift and competent response. Cardiopulmonary resuscitation (CPR) is a key lifesaving intervention for IHCA, but the frequency of CPR events and the decay of CPR skills and knowledge among healthcare professionals (HCPs) raise concerns.

**Methods and Results:** In a prospective quasi-experimental study with no control group, 263 registered nurses (121 in critical care units and 144 in general care units) were assessed for CPR skills and knowledge retention at various time intervals. The result showed that overall decay after six months are almost the same for both groups. Knowledge decay started for both groups after one month and for both groups the highest level of decay was reported after three months, however the decay pattern was quite different. General units mean differences showed sudden sharp decline on three months which was not reported with critical care nurses who showed steady decay reaching to six months.

**Conclusions:** In this study, it is noteworthy that nurses in critical care units exhibited consistent decay in skills and knowledge, while those in general care units demonstrated a progressive decline over time.

## 1. Introduction and Background

Cardiac arrest is defined as a sudden and unexpected stop of cardiac activity, resulting in a decrease in perfusion of blood to the vital organs, leading to an unresponsive victim with abnormal breathing and absence of signs of circulation (1). In-hospital cardiac arrest (IHCA) is the most urgent medical emergency and may lead to death when not recognized and treated with prompt initiation and appropriate interventions by competent providers (2). Generally, hospitals in the United States have been estimated to treat approximately 200,000 cardiac arrests annually (Bradley et al., 2019). This is an unpredictable event that often occurs without any prior warning, mandating quick and competent intervention.

Cardiopulmonary resuscitation (CPR) is an important life-saving medical intervention employed in response to cardiac arrest. It is designed to restore vital functions and maintain adequate blood flow to the essential organs when the heart suddenly stops beating effectively by chest compressions, ventilation and defibrillation (3). In general, research has demonstrated that when appropriately trained healthcare professionals (HCPs) administer CPR, it can lead to a decrease in in-hospital cardiac fatalities and related deaths (4).

The American Heart Association (AHA) plays a pivotal role in advancing the knowledge and practices related to cardiac resuscitation. As a preeminent organization in the realm of heart health, AHA has a rich history of conducting research, developing guidelines, dissemina t ing critical information pertaining to cardiac resuscitation science, and preparing HCPs to have the knowledge, skills, and self-efficacy necessary to act immediately in CPR (5). However, although the 2020 AHA guidelines recommend renewing certification every two years, HCPs have expressed anxiety about their required knowledge and CPR skills if they are exposed only infrequently to real CPR situations (4). Research has shown that infrequent CPR events might cause HCPs to lose confidence and consequently competence in responding to in-hospital cardiac arrest events due to deteriorating CPR knowledge and skills (4). Some studies suggested the need to teach CPR skills more frequently than the AHA biennial requirement (6–9). Lack of practice has been suggested as a possible explanation for the skills decay and suboptimal outcomes for patients requiring CPR (6,10).

Cardiopulmonary resuscitation skills decay overtime (11,12). This is also supported by multiple studies demonstrating that CPR skills decline too soon after training (13–15). Other studies have revealed that the compression rate and depth skills were retained at two months, three months, and six months (16,17). The most recent literature has revealed that CPR skills start to decay as early as two weeks, whereas knowledge retention declines on average between one month and six months after training (18). In conclusion, skills and knowledge retention may gradually decay among HCPs especially those who are not regularly exposed to CPR.

For this reason, the AHA has recently implemented the Resuscitation Quality Improvement (RQI) program as a means to uphold proficiency in CPR. This initiative focuses on ensuring that HCPs remain prepared for emergencies by engaging in frequent, low-volume training sessions. In order to maintain their Basic Life Support (BLS) or Advanced Certification through the RQI program, participants are required to exhibit their CPR proficiency on a manikin every 90 days. This increase in frequency of skill assessment is being implemented with the aim of avoiding any possible decay in HCPs’ skills (American Heart Association, 2017). However, it remains unclear what the optimal training time intervals are for different providers in various clinical specialties to maintain their skills to deliver the highest-quality CPR. The frequency with which HCPs encounter CPR cases depends on whether they work in critical care units or general care units. Critical care units typically have a higher frequency of CPR incidence due to the characteristics of their patients. Limited research has been conducted on the deterioration of CPR skills and knowledge among nurses in critical care and general care units. Consequently, the primary objective of this study is to assess the retention of CPR skills and knowledge among registered nurses (RNs) in critical care units in comparison to those in general care units.

## Materials and Method

### 2.1 Study design

A prospective quasi-experimental study without a control group was performed at different time intervals. Immediately after initial training, CPR skills and knowledge were assessed for all RNs (Ti), then a follow-up test performed at different timepoints of one month later (T1), three months later (T2), and six months later (T3) among the critical and general unit HCPs. Based on their respective working units, RNs were allocated to either the general unit group or the critical unit group. Within each group, RNs were randomly assigned to one of the three post-test measurement points T1, T2 and T3.

### 2.2 Recruitment & participants

We invited all the RNs (N= 300) from the critical care and general care units who were scheduled to attend a new Basic Life Support (BLS) course during the period from April 2022 to April 2023 to take part in this study. Of the 300 invited RNs, 275 agreed to participate in the study. However, only 265 of them completed the study and attended all the required tests, as 10 individuals voluntarily dropped out for personal reasons. To be eligible for inclusion, RNs had to work in a critical care or general care unit and have no physical limitations and/or any medical restrictions to performing CPR. All participants were provided with information regarding the research objective, study design, confidentiality assurance, and the voluntary nature of their participation, with the freedom to withdraw at any time without facing any consequences.

### 2.3 Data collection

Prior to the BLS training course, the participants were invited to the study, and consent was obtained. All the participants received the BLS training, and immediately after the course their evaluation was performed as an initial test (Ti). After that, participants were divided into critical care unit and general care unit groups based on their working location, and, anonymously, they were randomly assigned to have their post-tests at T1, T2, or T3. Participants were informed that the research team would arrange with their managers to invite them at a random time during their workday to perform the post-test. All these actions were taken to avoid any extra preparation for the post-test that may affect the research results. All courses were taught by experienced AHA-certified BLS instructors. The researchers obtained permission from the AHA to use their knowledge exam and the BLS skills evaluation checklist (2020 AHA guidelines). All measures were taken to secure the exam as per the training center policy. Meanwhile, in order to overcome any subjectivity of the skills evaluation, the researchers used the new high-fidelity CPR training manikins and devices during training and during post-training assessment, to obtain automated feedback regarding participants’ performance.

The accuracy of compression, breaths, and the ability to resume compressions within 10 seconds were evaluated during the second cycle of CPR using the skills reporter app. Subsequently, the arrival of the automated external defibrillator (AED) was simulated and assessed based on five criteria: powering up the AED, correct attachment of the pads, ensuring clarity for analysis, ensuring safety before delivering a shock, and safely delivering the shock. Finally, the participants’ ability to promptly resume compressions was assessed. It is important to note that the same AHA scenario was utilized for all participants, while the AHA written exam was administered to evaluate their knowledge in a very controlled environment to secure the exam.

On the days of the post-tests, participants were invited to do the test. Participants were given the same scenario and instructed by the instructor to act as they had been taught before. The circumstances were replicated to match the environment of the initial evaluation (Ti).

### 2.4 Ethical considerations

This study was approved by the King Hussein Cancer Center’s Institutional Review Board (number 22 KHCC 15), and the study purpose, methodology, and data management were explained in detail for the participants. Voluntary participation was assured for each participant, and they had the right to withdraw at any time, which would not affect their working status. Meanwhile, they received a unique code for their participation, and confidentiality and privacy were assured and maintained in accordance with the Declaration of Helsinki.

### 2.4 Data analysis

Data analysis was conducted using the statistical program SPSS version 29. A descriptive analysis was performed to examine the mean and standard deviation of the quantitative variables, while frequencies and percentages were calculated for the categorical variables. Statistical tests such as one-way ANOVA, paired t-test, and chi-square were employed to compare the initial test results with the post-test outcomes. Significance was determined using a p-value p < 0.05, indicating statistical significance.

## 3 Results

### 3.1 Sociodemographic characteristics

Table 1 displays the sociodemographic characteristics of the sample, divided into two main groups: the critical care unit group, consisting of 121 participants, and the general care unit group, consisting of 144 participants. There were no significant differences between these groups at the levels of gender or education. The majority of the total sample held a bachelor’s degree, and more than two-thirds of the participants were female. In terms of working experience, critical care unit nurses had an average of 4.53 years of experience, while those in the general care unit had an average of 6.47 years. The average age for the critical care unit group was 26.62 years, compared to 28.81 years for the general care unit group. Importantly, the results indicate significant differences in age and experience among the study sample.

**Table 1:**
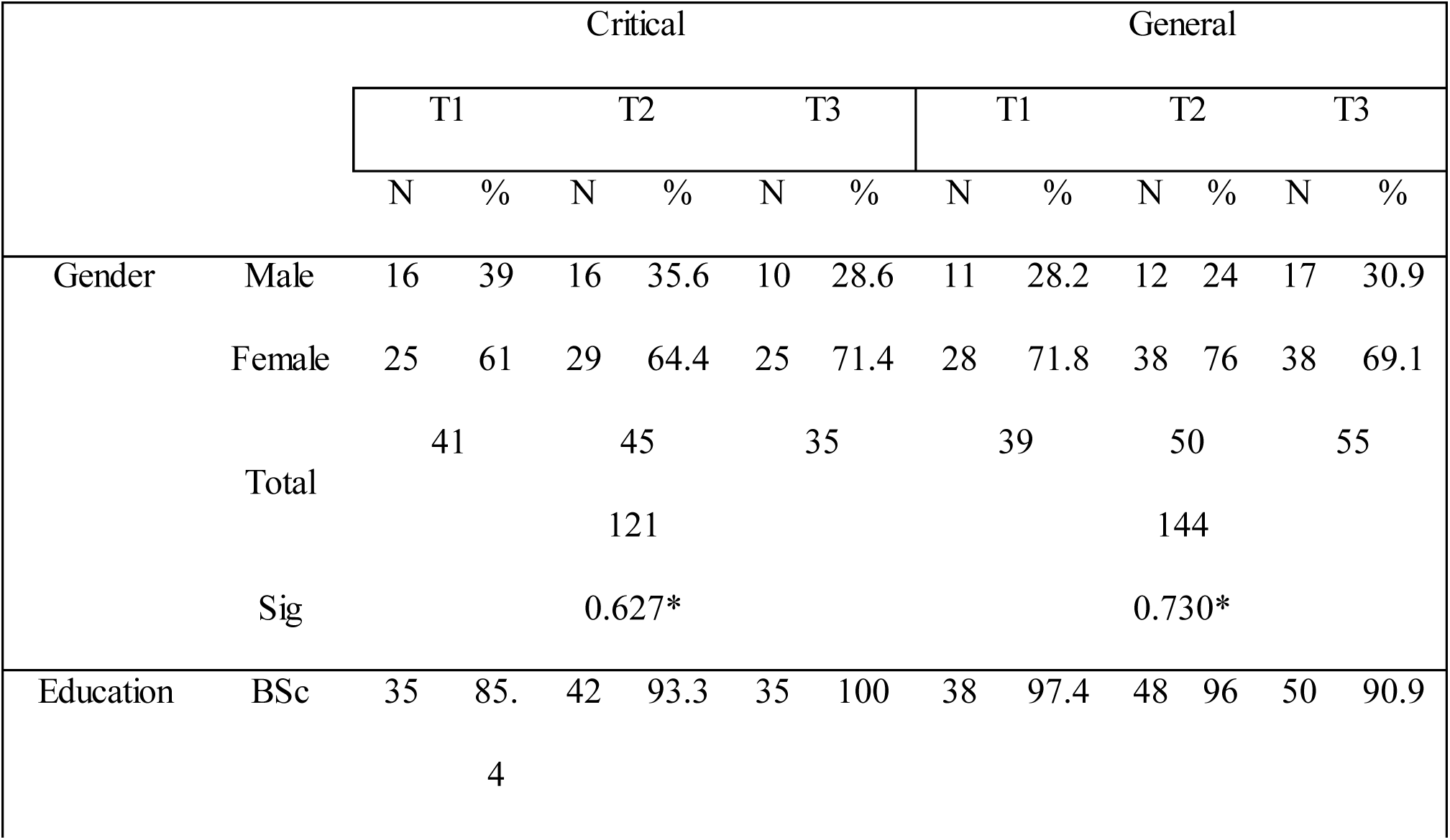
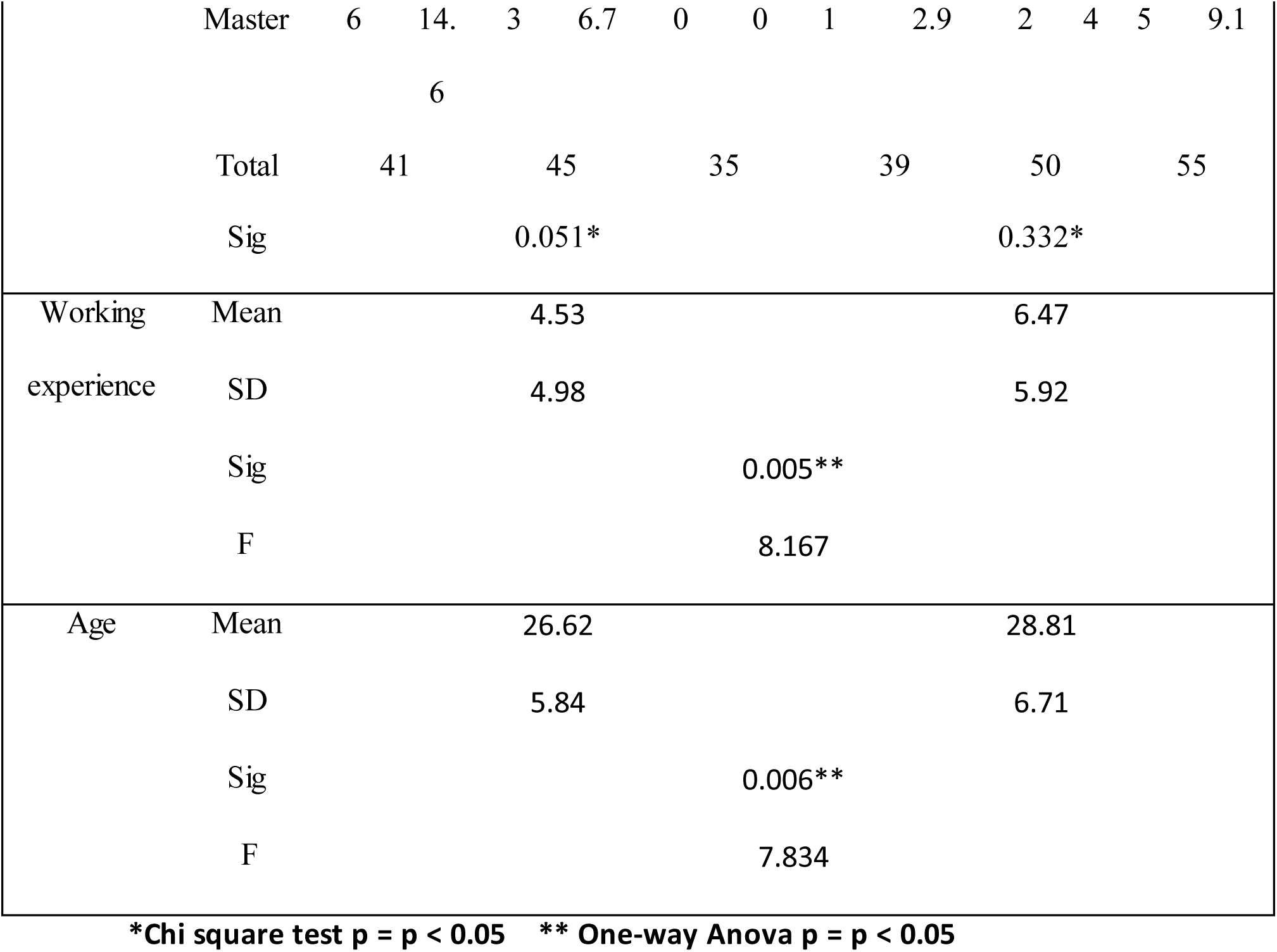
Socio-Demographic Characteristics of the Sample.

|  |  | Critical |  |  |  |  |  | General |  |  |  |  |  |
| --- | --- | --- | --- | --- | --- | --- | --- | --- | --- | --- | --- | --- | --- |
|  |  | T1 |  | T2 |  | T3 |  | T1 |  | T2 |  | T3 |  |
|  |  | N | % | N | % | N | % | N | % | N | % | N | % |
| Gender | Male | 16 | 39 | 16 | 35.6 | 10 | 28.6 | 11 | 28.2 | 12 | 24 | 17 | 30.9 |
|  | Female | 25 | 61 | 29 | 64.4 | 25 | 71.4 | 28 | 71.8 | 38 | 76 | 38 | 69.1 |
|  | Total | 41 |  | 45 |  | 35 |  | 39 |  | 50 |  | 55 |  |
|  |  | 121 |  |  |  |  |  | 144 |  |  |  |  |  |
|  | Sig | 0.627* |  |  |  |  |  | 0.730* |  |  |  |  |  |
| Education | BSc | 35 | 85. | 42 | 93.3 | 35 | 100 | 38 | 97.4 | 48 | 96 | 50 | 90.9 |
|  |  | 4 |  |  |  |  |  |  |  |  |  |  |  |
|  | Master | 6 | 14. | 3 | 6.7 | 0 | 0 | 1 | 2.9 | 2 | 4 | 5 | 9.1 |
|  |  |  | 6 |  |  |  |  |  |  |  |  |  |  |
|  | Total | 41 |  | 45 |  | 35 |  | 39 |  | 50 |  | 55 |  |
|  | Sig |  |  | 0.051* |  |  |  |  |  | 0.332* |  |  |  |
| Working | Mean |  |  | 4.53 |  |  |  |  |  | 6.47 |  |  |  |
| experience | SD |  |  | 4.98 |  |  |  |  |  | 5.92 |  |  |  |
|  | Sig |  |  |  |  |  |  | 0.005** |  |  |  |  |  |
|  | F |  |  |  |  |  |  | 8.167 |  |  |  |  |  |
| Age | Mean |  |  | 26.62 |  |  |  |  |  | 28.81 |  |  |  |
|  | SD |  |  | 5.84 |  |  |  |  |  | 6.71 |  |  |  |
|  | Sig |  |  |  |  |  |  | 0.006** |  |  |  |  |  |
|  | F |  |  |  |  |  |  | 7.834 |  |  |  |  |  |

**\*Chi square test $p = p < 0.05$ \*\* One-way Anova $p = p < 0.05$**

### 3.2 CPR knowledge evaluation

Table 2 shows the differences in knowledge competency mean scores at the level of groups and subgroups at Ti compared with T1, T2, and T3. For the critical care unit group, the total mean score of knowledge competency at Ti was 93.83, whereas it decreased to 83.83 at the post-test. Similarly, for the general care unit group, the mean score was 93.22 at Ti and the total mean score decreased to 82.54 at the post-test.

**Table 2:**
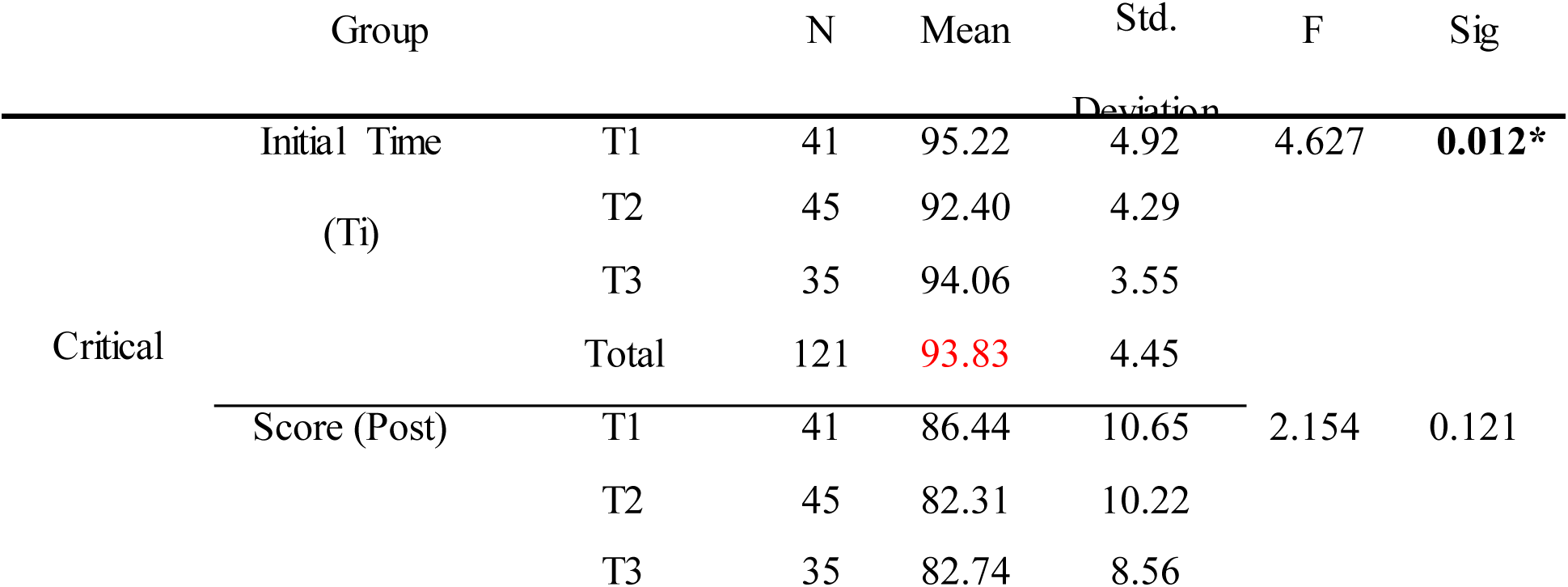
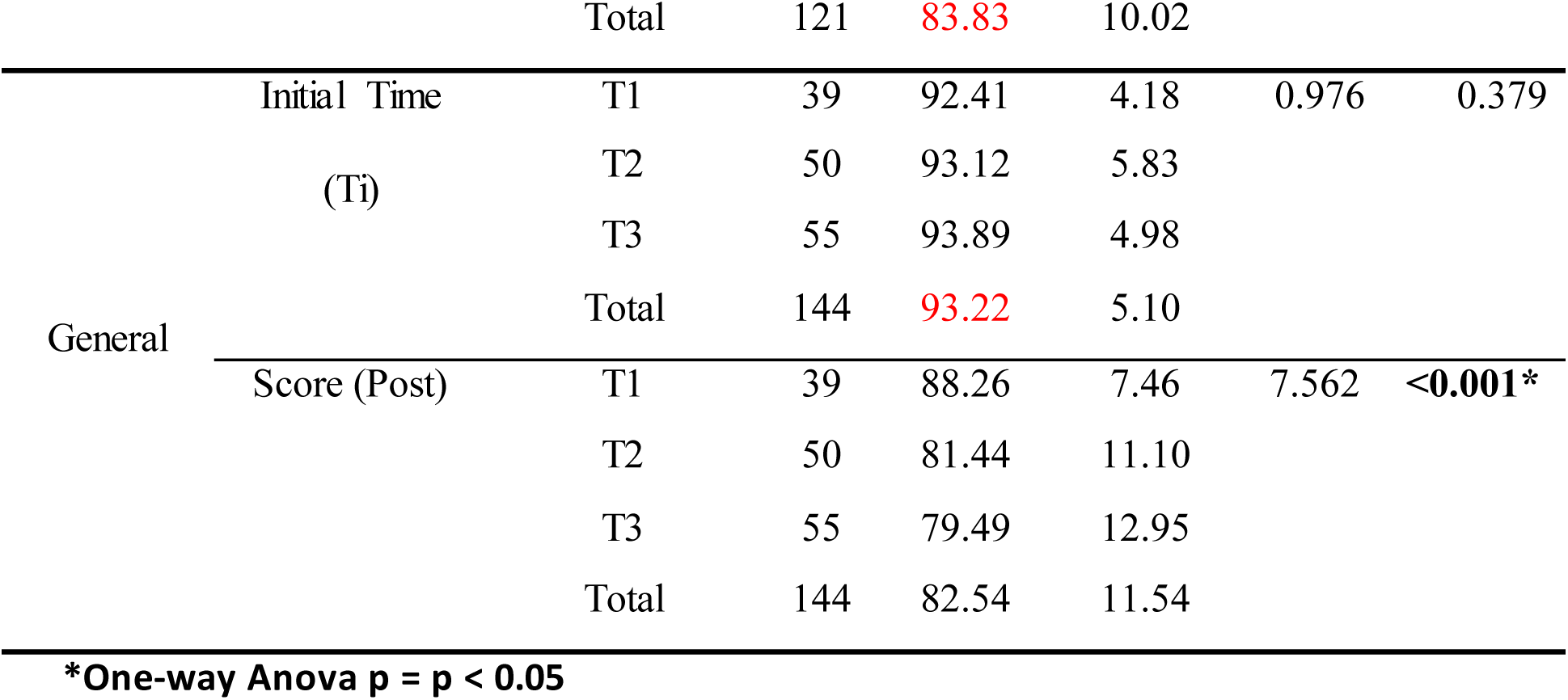
Knowledge Competency Differences at Different Time Points.

At the level of the subgroups’ knowledge competency mean score on Ti, Table 2 shows that the mean scores of the critical care units group at T1, T2, and T3 were 95.22, 92.40, and 94.06, respectively. In comparison, the general care unit group at T1, T2, and T3 had mean scores of 92.41, 93.12, and 93.89, respectively. At the post-test, the critical care unit group’s T1, T2, and T3 had mean scores of 86.44, 82.31, and 82.74, respectively. Similarly, the general care unit group’s T1, T2, and T3 had mean scores of 88.26, 81.44, and 79.49, respectively. Furthermore, Table 2 highlights significant differences in knowledge competency mean scores among the critical care unit group at the initial testing point Ti as (p value < 0.05), and the same significance was found among the general care unit group at post-test points as (p value < 0.05).

The Bonferroni post-hoc group comparisons conducted after the ANOVA analysis in Table 3 indicate that there were no significant differences between (T1 and T2) and (T1 and T3) within the critical care group. However, in the general care group, significant differences were observed between (T1 and T2) and (T1 and T3) as P value < 0.05.

**Table 3:**
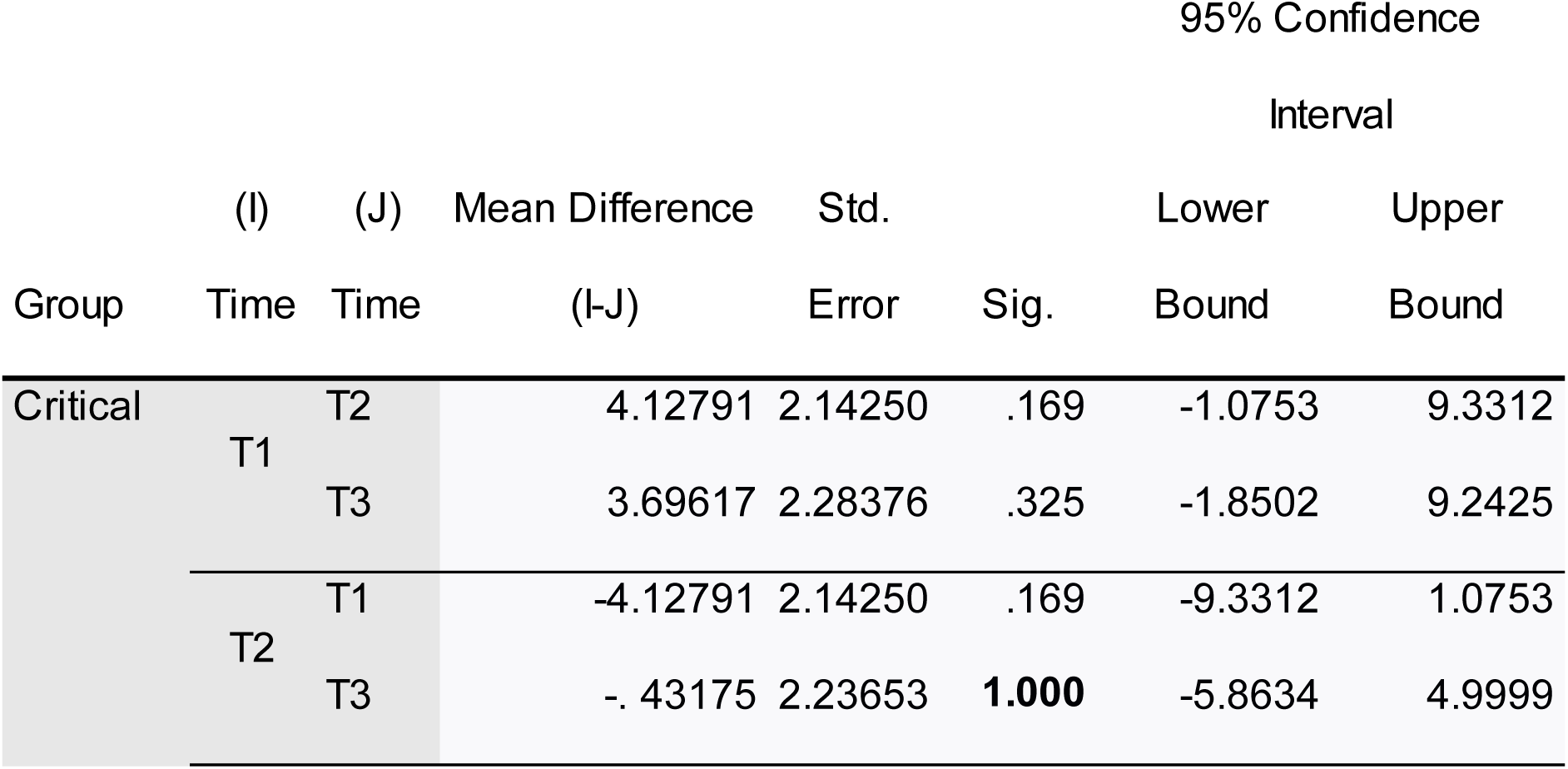
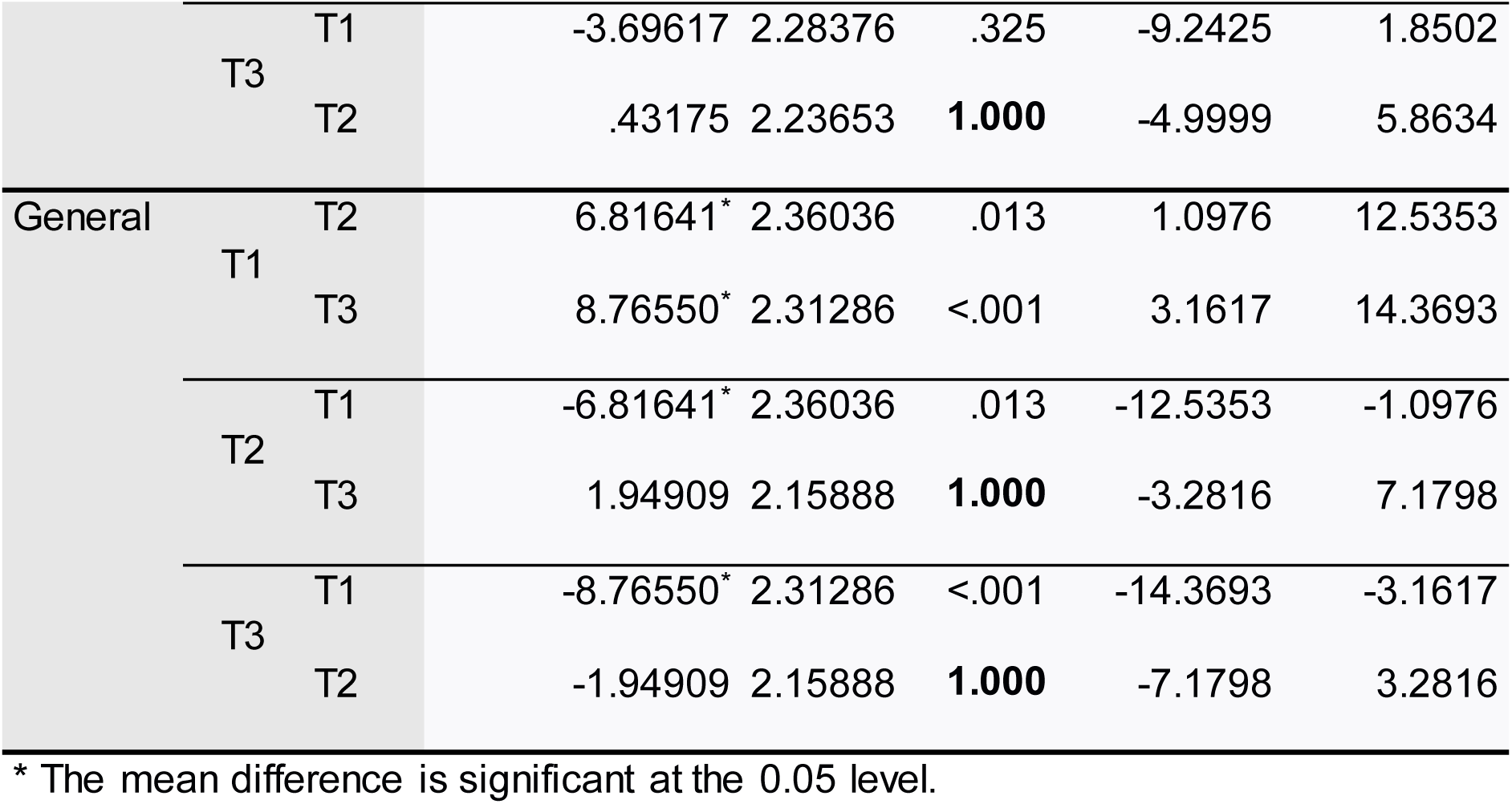
Multiple Comparison of Knowledge Mean Scores Between Total Subgroups.

### 3.3 CPR skills competency evaluation

Table 4 presents the skills competency evaluation final decision. At Ti all study participants passed the skills competency. In post-tests, within the critical care unit subgroups, the number of participants needing remediation (NR) was 34, 33, and 29 for T1, T2, and T3, respectively. In the general care unit subgroups’ post-tests, the overall count of participants needing remediation (NR) during T1, T2, and T3 was 29, 46, and 50, respectively. Moreover, the results show no significant differences among the critical care unit group at the time of comparing (T1 vs. T2), (T1 vs. T3), and (T2 vs. T3); while for the general care unit group, significant differences were found when comparing (T1 vs. T2) and (T1 vs. T3), with p values < 0.05. In addition to the paired test results, there are significant differences between the (T1, T2) and (T1, T3) of the general care unit group, with a P value of 0.05; whereas no significant differences were found at (T2, T3).

**Table 4:**
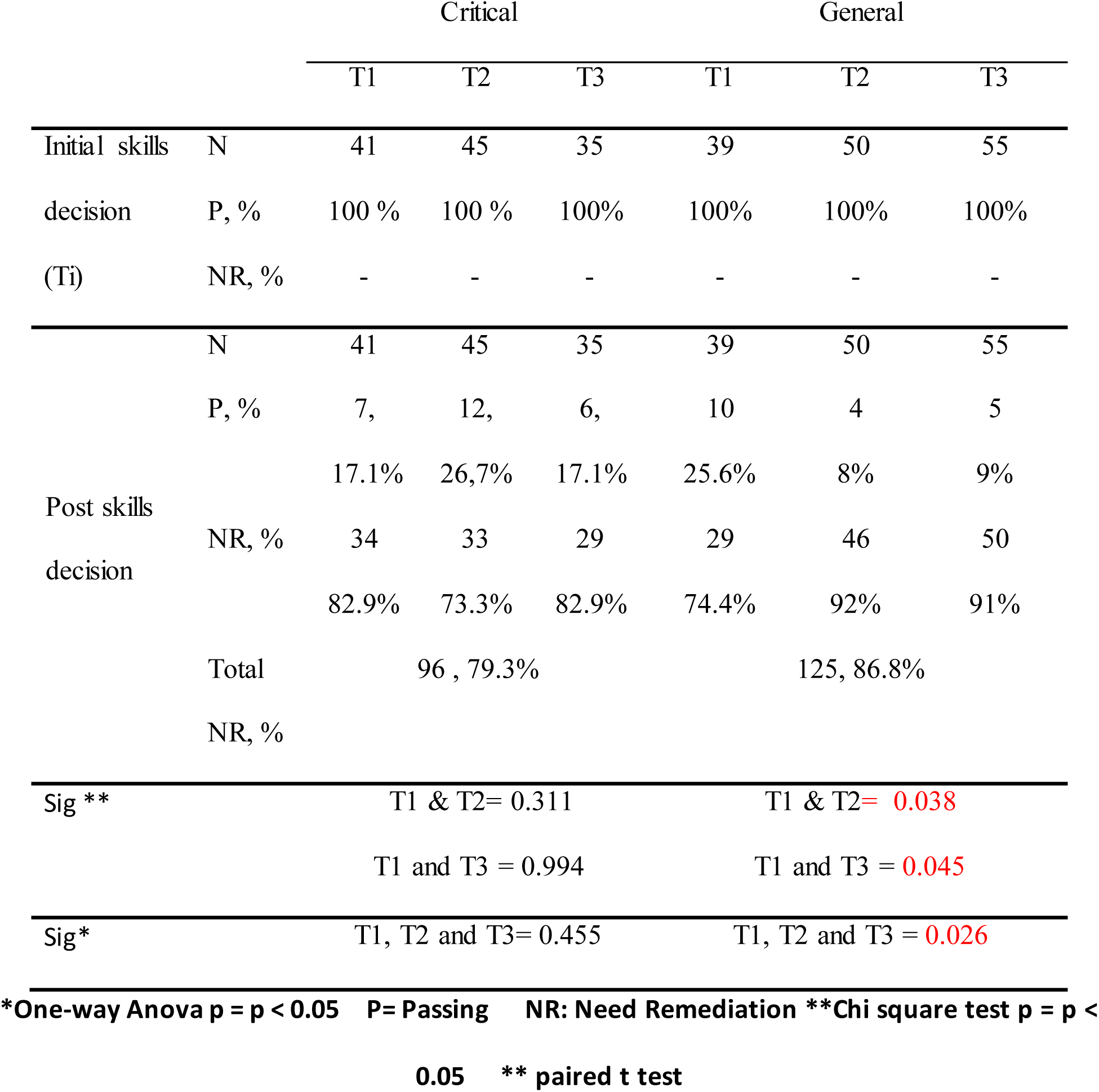
Final Decision of Skills Competency Evaluation at Level of Each Subgroup.

|  |  | Critical |  |  | General |  |  |
| --- | --- | --- | --- | --- | --- | --- | --- |
|  |  | T1 | T2 | T3 | T1 | T2 | T3 |
| Initial skills | N | 41 | 45 | 35 | 39 | 50 | 55 |
| decision | P, % | 100 % | 100 % | 100% | 100% | 100% | 100% |
| (Ti) | NR, % | - | - | - | - | - | - |
| Post skills<br>decision | N | 41 | 45 | 35 | 39 | 50 | 55 |
|  | P, % | 7, | 12, | 6, | 10 | 4 | 5 |
|  |  | 17.1% | 26,7% | 17.1% | 25.6% | 8% | 9% |
|  | NR, % | 34 | 33 | 29 | 29 | 46 | 50 |
|  |  | 82.9% | 73.3% | 82.9% | 74.4% | 92% | 91% |
|  | Total | 96 , 79.3% |  |  | 125, 86.8% |  |  |
|  | NR, % |  |  |  |  |  |  |
| Sig ** |  | T1 & T2= 0.311 |  |  | T1 & T2= **0.038** |  |  |
|  |  | T1 and T3 = 0.994 |  |  | T1 and T3 = **0.045** |  |  |
| Sig* |  | T1, T2 and T3= 0.455 |  |  | T1, T2 and T3 = **0.026** |  |  |

**\*One-way Anova p = p < 0.05 P= Passing NR: Need Remediation \*\*Chi square test p = p <**
**0.05 \*\* paired t test**

## 4. Discussion

The purpose of this study was to determine the threshold at which the cardiopulmonary resuscitation competency among registered nurses declines to a level that no longer meets the latest standards set by the American Heart Association (AHA) in 2020. Research suggests that healthcare professionals, including RNs, may experience a decay in BLS knowledge and skills over time in the absence of consistent practice (10,19). Factors such as infrequent exposure to emergencies, lack of practice opportunities, and high workloads, can contribute to skills decay. This deterioration in skills and knowledge can result in suboptimal professional performance when caring for cardiac arrest patients, ultimately leading to decreased patient survival rates (10). However, none of the reviewed literature reported the differences between RNs in different care settings.

### 4.1 CPR knowledge and skills after initial training

Notably, during the initial assessment (Ti), the critical care unit group achieved a higher mean score of knowledge competency than the general care unit group. Furthermore, when we examine the total mean knowledge competency score at the Ti level and after re-assessment (post-score), it becomes evident that the critical care unit team displayed smaller mean score discrepancies compared to the general care unit team. There are several possible explanations for these results; the frequency of CPR incidence in critical care areas may have played a role in maintaining the knowledge and skills of HCPs in that unit. In addition, critical care settings often experience more acute patient care situations, which may require HCPs to be exposed to some CPR-associated skills on a more regular basis. This constant exposure to real-life scenarios could contribute to a better retention of CPR knowledge and skills over time.

Also, the study noted that the RNs in the critical care unit were generally younger than those in the general care unit. Since younger individuals tend to have more agile cognitive abilities, making it easier for them to comprehend, process, and remember new information, and indeed numerous studies have discussed and provided explanations regarding the impact of age, the nature of critical care work, and the frequency of CPR on the acquisition and retention of informat io n, this aligns with the findings of this study (Borovnik Lesjak et al., 2022; De Araujo et al., 2021; Guskuma et al., 2019; Rajeswaran et al., 2018). In terms of skills competency, the findings revealed that all participants from both groups were able to successfully pass the skills evaluation immediately after completing their CPR training. This outcome aligns with prior research in the field, which has consistently emphasized the effectiveness of CPR training programs that employ high-fidelity manikins and follow standardized training methodologies in enhancing CPR skills competency (23,19,24). These collective findings underscore the importance of these training approaches in equipping RNs with the necessary skills to save lives through CPR interventions. However, the question is: for how long can this competency be maintained?

### 4.2 CPR knowledge and skills decay at different time intervals

The results of this study highlight that participants from both groups demonstrated knowledge and skills decay after initial training, as has already been documented in the literature (10,19,24–26). Many participants did not exhibit optimal CPR knowledge and skills over different time intervals after initial training. In fact, many participants were remediated on essential CPR skills; this was also reported in similar studies (27). However, in terms of skills competency, it was revealed that the critical care unit group had a lower percentage of remediation compared to the general care unit group. This finding, as explained earlier, could be attributed to factors such as age, acuity of patient cases, and the frequency of CPR occurrences, which may contribute to a more sustained retention of CPR skills.

For critical care unit nurses, the knowledge domain over the different time intervals showed a sudden drop in decay one month after the initial training, and then the level of knowledge was almost maintained afterward at the three- and six-month time intervals. The same trend was observed for the skills competency, and this is very remarkable. This could be understandable in terms of knowledge retention; however, it may also help clinical educators to plan a complementary skills training dose (‘refresher’) at this point. More research may shed more light on this matter.

For the general care unit nurses, the case was quite different. The nurses had the same major drop in the knowledge part after the first month, while they then maintained minor progressive decay afterward at the three- and six-month time intervals. The same trend was also noticed with the skills competency. The progressive decay may be related to the lower acuity in patient care cases, which have fewer cases of CPR, and the linked skills competencies.

Critical care unit nurses showed less overall knowledge and skills decay after six months than general care unit nurses and this was discussed and justified above. Both groups had the same level of knowledge and skills decay after one month, which is remarkable because majority of the available reviewed literature indicated that the decay may start happening at around 3 months. Moreover, the differences in knowledge and skills decay patterns among nurses working in critical care and general care units was not reported before, and therefore this is unique to this study. It is extraordinary to find out in this study that nurses in critical care units had steady and constant decay over time, while nurses in general units had progressive decay overtime, and this may need further investigation. This is very crucial for clinical educators to plan and design training refreshers for both care areas based on the differences in knowledge and skills decay patterns.

## Conclusion

In conclusion, this study aimed to identify the threshold at which CPR competency among registered nurses no longer meets the 2020 American Heart Association standards. The findings reveal distinct patterns of knowledge and skills decay between critical care and general care unit nurses. Critical care unit nurses displayed steadier knowledge and skills retention, potentially attributed to higher exposure to CPR scenarios and they are younger in age. In contrast, general care unit nurses experienced progressive decay, likely due to lower acuity cases. Remarkably, both groups exhibited significant knowledge and skills decay after one month, differing from previous literature suggesting decay at around three months. This novel insight underscores the need for tailored training refreshers based on the observed decay patterns, calling for further investiga t ion in this critical area for the benefit of patient outcomes and healthcare education.

## Data Availability

All data referred to in the manuscript is available upon reques.

## References

1. Sharabi AF, Singh A. Cardiopulmonary Arrest in Adults. In: StatPearls [Internet]. Treasure Island (FL): StatPearls Publishing; 2023 [cited 2023 Jun 13]. Available from: http://www.ncbi.nlm.nih.gov/books/NBK563231/

2. Khera R, Tang Y, Link MS, Krumholz HM, Girotra S, Chan PS, et al. Association Between Hospital Recognition for Resuscitation Guideline Adherence and Rates of Survival for In-Hospital Cardiac Arrest. Circ Cardiovasc Qual Outcomes. 2019 Mar;12(3):e005429.

3. Giza DE, Rodrigo M, Crommett J, Botz G, Ewer MS, Iliescu G, et al. Impact of cardiopulmonary resuscitation (CPR) on the survival of patients with cancer: DNR before or after cardiac arrest? J Clin Oncol. 2018 Dec 1;36(34_suppl):71–71.

4. Rajeswaran L, Cox M, Moeng S, Tsima BM. Assessment of nurses’ cardiopulmonary resuscitation knowledge and skills within three district hospitals in Botswana. Afr J Prim Health Care Fam Med [Internet]. 2018 Apr 12 [cited 2023 Jun 13];10(1). Available from: https://phcfm.org/index.php/phcfm/article/view/1633

5. Berg KM, Cheng A, Panchal AR, Topjian AA, Aziz K, Bhanji F, et al. Part 7: Systems of Care: 2020 American Heart Association Guidelines for Cardiopulmonary Resuscitation and Emergency Cardiovascular Care. Circulation [Internet]. 2020 Oct 20 [cited 2023 Jun 13];142(16_suppl_2). Available from: https://www.ahajournals.org/doi/10.1161/CIR.0000000000000899

6. Skoblar R. Cardiopulmonary Resuscitation (CPR) Skill Refreshers for Nurses: An Evidence-Based Project. Fairleigh Dickinson University; 2020.

7. Lin Y, Cheng A, Grant VJ, Currie GR, Hecker KG. Improving CPR quality with distributed practice and real-time feedback in pediatric healthcare providers – A randomized controlled trial. Resuscitation. 2018 Sep;130:6–12.

8. Saramma PP, Raj LS, Dash PK, Sarma PS. Assessment of long-term impact of formal certified cardiopulmonary resuscitation training program among nurses. Indian J Crit Care Med Peer-Rev Off Publ Indian Soc Crit Care Med. 2016 Apr;20(4):226–32.

9. Cheng A, Brown LL, Duff JP, Davidson J, Overly F, Tofil NM, et al. Improving Cardiopulmonary Resuscitation With a CPR Feedback Device and Refresher Simulations (CPR CARES Study): A Randomized Clinical Trial. JAMA Pediatr. 2015 Feb 1;169(2):137.

10. Cheng A, Nadkarni VM, Mancini MB, Hunt EA, Sinz EH, Merchant RM, et al. Resuscitation Education Science: Educational Strategies to Improve Outcomes From Cardiac Arrest: A Scientific Statement From the American Heart Association. Circulation [Internet]. 2018 Aug 7 [cited 2023 Jul 4];138(6). Available from: https://www.ahajournals.org/doi/10.1161/CIR.0000000000000583

11. Schmitz GR, McNeilly C, Hoebee S, Blutinger E, Fernandez J, Kang C, et al. Cardiopulmonary resuscitation and skill retention in emergency physicians. Am J Emerg Med. 2021 Mar;41:179–83.

12. Kim YJ, Cho Y, Cho GC, Ji HK, Han SY, Lee JH. Retention of cardiopulmonary resuscitation skills after hands-only training versus conventional training in novices: a randomized controlled trial. Clin Exp Emerg Med. 2017 Jun 30;4(2):88–93.

13. Charlier N, Van Der Stock L, Iserbyt P. Comparing student nurse knowledge and performance of basic life support algorithm actions: An observational post-retention test design study. Nurse Educ Pract. 2020 Feb;43:102714.

14. Anderson R, Sebaldt A, Lin Y, Cheng A. Optimal training frequency for acquisition and retention of high-quality CPR skills: A randomized trial. Resuscitation. 2019 Feb;135:153–61.

15. Mokhtari Nori J, Saghafinia M, Kalantar Motamedi MH, Khademol Hosseini SM. CPR Training for Nurses: How often Is It Necessary? Iran Red Crescent Med J. 2012 Feb;14(2):104– 7.

16. Hong C, Hwang S, Lee K, Kim Y, Ha Y, Park S. Metronome vs. Popular Song: A Comparison of Long-Term Retention of Chest Compression Skills after Layperson Training for Cardiopulmonary Resuscitation. Hong Kong J Emerg Med. 2016 May;23(3):145–52.

17. Bobrow BJ, Vadeboncoeur TF, Spaite DW, Potts J, Denninghoff K, Chikani V, et al. The Effectiveness of Ultrabrief and Brief Educational Videos for Training Lay Responders in Hands-Only Cardiopulmonary Resuscitation: Implications for the Future of Citizen Cardiopulmonary Resuscitation Training. Circ Cardiovasc Qual Outcomes. 2011 Mar;4(2):220–6.

18. Ahmed S, Ismail I, Lee K, Lim P. Systematic review on knowledge and skills level among nurses following cardiopulmonary resuscitation (cpr) training [Internet]. In Review; 2021 Oct [cited 2023 Jun 13]. Available from: https://www.researchsquare.com/article/rs-951043/v1

19. Aqel AA, Ahmad MM. High-Fidelity Simulation Effects on CPR Knowledge, Skills, Acquisition, and Retention in Nursing Students: HFS and CPR Knowledge and Skills. Worldviews Evid Based Nurs. 2014 Dec;11(6):394–400.

20. Borovnik Lesjak V, Šorgo A, Strnad M. Retention of Knowledge and Ski **l**s After a Basic Life Support Course for Schoolchildren: A Prospective Study. Inq J Health Care Organ Provis Financ. 2022 Jan;59:004695802210987.

21. De Araujo NR, Moretti MA, De Araújo RA, Chagas ACP. Retention of Knowledge and Skills in Cardiopulmonary Resuscitation Among Clinical and Intensive Care Professionals After Educational Intervention [Internet]. In Review; 2021 Aug [cited 2023 Jul 5]. Available from: https://www.researchsquare.com/article/rs-725603/v1

22. Guskuma EM, Lopes MCBT, Piacezzi LHV, Okuno MFP, Batista REA, Campanharo CRV. Conhecimento da equipe de enfermagem sobre ressuscitação cardiopulmonar em um hospital universitário. Rev Eletrônica Enferm [Internet]. 2019 Dec 31 [cited 2023 Jul 4];21. Available from: https://www.revistas.ufg.br/fen/article/view/52253

23. Tosif S, Jatobatu A, Maepioh A, Gray A, Sobel H, Mannava P, et al. Healthcare worker knowledge and skills following coaching in WHO early essential newborn care program in the Solomon Islands: a prospective multi-site cohort study. BMC Pregnancy Childbirth. 2020 Dec;20(1):84.

24. Ackermann AD. Investigation of Learning Outcomes for the Acquisition and Retention of CPR Knowledge and Skills Learned with the Use of High-Fidelity Simulation. Clin Simul Nurs. 2009 Nov;5(6):e213–22.

25. Bradley SM, Zhou Y, Ramachandran SK, Engoren M, Donnino M, Girotra S. Retrospective cohort study of hospital variation in airway management during in-hospital cardiac arrest and the association with patient survival: insights from Get With The Guidelines-Resuscitation. Crit Care. 2019 Dec;23(1):158.

26. Kleinman ME, Goldberger ZD, Rea T, Swor RA, Bobrow BJ, Brennan EE, et al. 2017 American Heart Association Focused Update on Adult Basic Life Support and Cardiopulmonary Resuscitation Quality: An Update to the American Heart Association Guidelines for Cardiopulmonary Resuscitation and Emergency Cardiovascular Care. Circulation [Internet]. 2018 Jan 2 [cited 2023 Jul 10];137(1). Available from: https://www.ahajournals.org/doi/10.1161/CIR.0000000000000539

27. Madden C. Undergraduate nursing students’ acquisition and retention of CPR knowledge and skills. Nurse Educ Today. 2006 Apr;26(3):218–27.

